# Interrupted time series analyses to assess the impact of alcohol control policy on socioeconomic inequalities in mortality in Lithuania: a study protocol

**DOI:** 10.1101/2021.05.10.21256944

**Authors:** Jakob Manthey, Domantas Jasilionis, Huan Jiang, Olga Meščeriakova-Veliulienė, Janina Petkevičienė, Ričardas Radišauskas, Jürgen Rehm, Mindaugas Štelemėkas

## Abstract

**Introduction:** Alcohol use is a major risk factor for mortality. Previous studies suggest that the alcohol-attributable mortality burden is higher in lower socioeconomic strata. This project will test the hypothesis, that the 2017 increase of alcohol excise taxes for beer and wine, which was linked to lower all-cause mortality rates in previous analyses, will reduce socioeconomic mortality inequalities.

**Methods and analysis:** Data on all causes of deaths will be obtained from Statistics Lithuania. Record linkage will be implemented using personal identifiers combining data from 1) the 2011 whole-population census, 2) death records between March 1, 2011 (census date) and December 31, 2019, and 3) emigration records, for individuals aged 30 to 70 years. The analyses will be performed separately for all-cause and for alcohol-attributable deaths. Monthly age-standardized mortality rates will be calculated by sex, education, and three measures of socioeconomic status. Inequalities in mortality will be assessed using absolute and relative indicators between low and high SES groups. We will perform interrupted time series analyses, and test the impact of the 2017 rise in alcohol excise taxation using generalized additive mixed models. In these models, we will control for secular trends for economic development.

**Ethics and dissemination:** This work is part of project grant 1R01AA028224-01 by the National Institute on Alcohol Abuse and Alcoholism. It has been granted research ethics approval 050/2020 by CAMH Research Ethics Board on April 17, 2020, renewed on March 30, 2021.

**Strengths and limitations of this study:**

- Census-linked mortality data will cover the entire population of Lithuania aged 30 to 70 years
- Three different definitions of socioeconomic groupings will allow for a comprehensive description of mortality inequalities in Lithuania between 2011 and 2019.
- The results will indicate if and to what extent an increase of alcohol excise taxes may be effective for reducing of health inequalities, thus closing the evidence gap of modelling studies predicting alcohol taxation to be an effective tool to reduce health inequalities.
- Interpretation of the effects will depend on identifying relevant confounders.
- An important limitation of census-linked studies is that the socioeconomic grouping is fixed at the census baseline.

## Introduction

Health inequalities have been internationally recognized as a major public health problem which persists even in rather egalitarian high-income countries in the North of Europe.[1] National and international policies have indicated the importance of health inequalities and proposing strategies towards reducing inequalities.[2, 3]

Health inequalities are most commonly frequently defined by mortality and life expectancy differences between groups with low and high socioeconomic status (SES). Depending on the definition employed, diverging trends can be observed in Europe.[4] Using absolute indicators of health inequality, substantial reductions could be observed in many European countries between 1990 and 2012, whereas in relative terms, the picture was mixed.[5]

Lithuania has been identified as one of the European countries with high levels of health inequalities [5]. For example, studies show that mortality rates have substantially increased between 1990-94 and 2005-09 among low-educated persons, starkly contrasting decreasing trends in mortality among low-educated persons from ten other European locations.[6] Comparing data from 2001 and 2014, both absolute and relative mortality gaps between low- and high educated persons were found to have widened in Lithuania.[7] In this period, the socioeconomic difference in all-cause mortality rates increased among men and women by about one third. A more refined set of analyses using data of life expectancy at age 30 between 2001 and 2014 identified breaking points in the trajectories of health inequalities in Lithuania.[8] The results suggest that lower educated males did not show any improvements, while life expectancy among higher educated men increased starting in 2006, suggesting a widening of health inequalities between 2006 and 2014. For women, a similar pattern was observed.

Several potential contributors to health inequalities have been identified for Lithuania,[9] and alcohol consumption and the attributable harm is one of them. Data for 2011-2015 show that deaths from alcohol-attributable diseases were 4.6 times more likely among low-educated persons, as compared to the high-educated counterparts.[10] This contrasts to 2.1 times higher all-cause mortality rates among low vs. high-educated Lithuanians in this period. Given the high levels of per capita consumption and alcohol-attributable disease burden,[11-13] alcohol is likely to be on key driver for health inequalities in Lithuania.

Since 2008, a number of effective alcohol policies have been enacted in Lithuania, ranging from bans on advertising, increasing drink-driving penalties, reducing the availability of alcohol by banning sales at petrol stations or restricting sales hours, and increasing taxes for alcoholic beverages.[14] Using time series analyses, the implementation of alcohol policies haven been linked to a reduction in alcohol-attributable motor vehicle accidents and injuries [15], as well as to declines in all-cause mortality.[16] The most robust evidence for reduction in mortality has converged for an increase in alcohol excise taxes (111-112% for wines and beer; 23% for ethyl alcohol), implemented on 1 March 2017.[14, 17]

While the evidence for reducing overall harm through alcohol control policies is strong, studies on their impact on health inequalities are sparse. Following micro-economic theories, it can be hypothesized that the impact of price increases on purchasing behavior will be greater among people with less disposable income, making taxation-based policies an appealing option to reduce health inequalities.[18] Accordingly, raising the floor price of alcoholic beverages in Scotland has resulted in stark reductions in alcohol purchases among more deprived households.[19]

In this article, we present a study protocol for assessing a) the trajectory of inequalities in mortality in Lithuania since the year 2012, and b) the potential impact of the recently introduced increase in alcohol excise taxes in Lithuania on changes in the magnitude of absolute and relative mortality inequalities by selected characteristics of socioeconomic status.

## Methods and analysis

### Preparing the data

The target population is the population aged 30 to 70 participating in the 2011 census in Lithuania. Linking census data with death records, the target population will be followed up for a total of 106 months, from March 2011 to December 2019. The monthly time series analyses will be restricted to the period from January 2012 to December 2019.

The census-linked mortality data includes information on sex, date of birth, educational attainment at census, economic activity, occupation, date of death, cause of death (4 digit ICD-10 code), place of residence (urban, rural), and emigration details (status and date of emigration). Information on emigration is needed to calculate person-months of exposure.

For calculating monthly age-standardized mortality rates, a breakup of the number of deaths (nominator) and person-months (denominator) for each sex-age-SES group and for each month will be calculated. For the nominator, the number of deaths can be directly calculated. For the denominator, we will calculate the average yearly person-months and obtain monthly estimates by first dividing the yearly estimates by 12 and second linearly interpolating between years. Persons emigrating from Lithuania and not returning until the end of the observational period will be included into the calculations of person months of exposure until the emigration date, whereas persons immigrating to Lithuania after the census are excluded from the analyses. As the assignment of persons to age groups will change over time, the allocation of nominator and denominator to the correct age bands will be required. We will use the ‘age-of-death’ format as described by Mackenbach and colleagues,[20] i.e., allocating deaths and person-years to the *current* age band. For person-years, this will represent the longest period of the year at a specific age (e.g. 39 if the person turned 39 in February, or 42 if the person turned 43 in October).

To account for differences in age-structure within sociodemopraphic groupings, we will calculate monthly sex- and SES-stratified age-standardized mortality rates, using the mid-point of the study period as standard population.

### Defining dependent variables

As dependent variables, we will calculate absolute and relative indicators of mortality inequalities. Specifically, this will be a) the absolute difference in age-standardized mortality rates between low and high SES groups, and b) the ratio of age-standardized mortality rates between low and high SES groups.

There will be two sets of dependent variables, defined by diagnostic grouping: 1) all-cause mortality, 2) an indicator for alcohol-attributable diseases (ICD-10 diagnostic codes: K70, K74, X45, T51, F10, G31.2, G62.1, I42.6, K29.2, K86.0, K85.2). For liver disease, we use the wider categories of K70 and K74, as there had been several coding rule changes for K70 in Lithuania during the time period examined. The combined indicator may result in a more consistent indicator even though not all of the cases in K74 category are alcohol-attributable.

The definition of SES groups will be based on a three-level educational attainment recorded at the 2011 census, as this is the most time-invariant SES measure available. In line with ISCED the definition, the highest educational attainment will be grouped into the following categories:

- ISCED 0-2 (low educational achievement): vocational school after graduating from a lower secondary school; vocational school without completion of a lower secondary school and acquired a lower secondary education together with a profession; vocational school without completion of a lower secondary school; a lower secondary school; (unfinished) primary school; literate (no schooling); illiterate; unknown.
- ISCED 3-4 (middle educational achievement): professional college; special secondary school; upper secondary school, gymnasium; vocational school after graduating from an upper secondary school; vocational school after graduating from a lower second school and acquired an upper secondary education together with the profession.
- ISCED 5-8 (high educational achievement): doctoral studies; university; college.

In additional analyses, we will refer to two alternative SES groupings: First, for manual vs. non-manual and other workers (restricted to the population employed at time of census and defined using the 2008 International Standard Classification of Occupations [21]), and second, for the urban vs. rural population.

Thus, there will be twelve dependent variables in total: two mortality inequality indicators (absolute and relative) for two mortality groupings (all-cause and alcohol) and three different sociodemographic groupings (education, occupational status, urban-rural).

### Independent variables

As independent variables, we will consider to include a secular trend variable (linear, quadratic, cubic), unemployment rates, income, and gross-domestic product. These variables will be used to account for possible confounders and to make the time series stationary. Further, we will include a binary variable for examining the 2017 policy, coded “0” in all months prior to March 2017 and “1” in all months March 2017 onwards, assuming an abrupt and persistent level change in the dependent variable. Based on visual inspections of the time series, slope changes and lagged effects will be considered.

### Statistical analyses

We will perform interrupted time series analyses, testing the impact of the 2017 rise in alcohol excise taxation using generalized additive mixed models (following recommendations of Beard and colleagues:[22]; previous applications on Lithuanian mortality data:[15-17]). For this, we will first build baseline models with control variables (secular trend, unemployment, GDP) and then include the policy variable to test for an immediate level change in the dependent variable.

Generalized additive mixed models allow to correct for autocorrelation by adding autoregressive or moving average terms. Seasonality will be accounted for using smoothing splines. Lastly, the residuals in all models will be checked for stationarity using augmented Dickey-Fuller tests.

## Ethics and dissemination

The proposed analyses will be carried out as part of a larger, NIAAA-funded project on evaluating alcohol control policies in Lithuania (grant no. 1R01AA028224-01). The project has undergone formal ethical review from the Research Ethics Board of the Centre for Addiction and Mental Health (CAMH REB), Toronto, Canada (REB decision letter no. 050/2020). Further, the Lithuanian Bioethics Committee has confirmed that handling of individual-level census and mortality data is not subject to the requirements of the Law on the Ethics of Biomedical Research in Lithuania (certificate no. 6B-17-91).

The census-linked mortality data will be obtained from Statistics Lithuania. All procedures involving individual record linkages were performed at Statistics Lithuania following the rules of data confidentiality and by employees having permission to work with confidential data. For the purpose of the proposed study, individual-level anonymized data are aggregated to calculate monthly age-standardized mortality rates. These mortality rates will constitute the base for the statistical analyses outlined in this study protocol and will not allow to identify natural persons.

As with previous studies of the above mentioned project,[16] the underlying aggregated time series of mortality inequalities and the statistical code will be published together with the results. This will facilitate further analyses of the same data and the adaptation of the proposed methods to other data.

### Patient and Public Involvement

Patients and the public were not involved in conceptualizing the proposed study.

## Discussion

It has been recognized that health inequalities serve as major barrier for both the future health improvements and social development.[23] While the contribution of lifestyle risk factors, such as smoking and alcohol use, to health inequalities are well known,[24, 25] the evidence base of alcohol- and smoking control policies to reduce these health inequalities remains scarce.

High levels of alcohol consumption and a comparably large attributable disease burden globally [11] and particularly in Europe [13] require effective public health interventions, such as increasing the retail price of alcoholic beverages.[26, 27] The more than 100% excise tax increase for beer and wine implemented in March 2017 in Lithuania has been linked to a reduction of all-cause mortality.[16] Extending the evaluation of this and other alcohol control policies to health inequalities, as proposed in this study, is key to assess possible unintended negative consequences, such as widening the gap between people with low and high socioeconomic status. As such, it will be one of the first studies using empirical data to evaluate the impact of alcohol control policies on health inequalities (modelling studies:[18, 28]).

This study will also serve as benchmark for additional studies on health inequalities in Lithuania and elsewhere. Being embedded in a multi-country NIAAA-funded project to evaluate the effects of alcohol policies in Lithuania and other Baltic countries, we expect this study to serve as benchmark for further applications, e.g. to evaluate other alcohol control policies (e.g.,[29]), to make age-specific analyses, or to use other outcomes, such as hospitalizations.

This study has several strenghts. First, we use census-linked mortality data covering the entire national population of Lithuania. Census-linkage of death records is one of the few possibilities to study the socioeconomic variation in mortality in a given country. As such data remain scarce in the Central and Eastern European region, our study will provide important insights also for neighbouring countries. Second, the study uses both relative and absolute indicators of socioeconomic inequalities in mortality, thus, providing a complete assessment of mortality inequalities in Lithuania.[4] Third, we test our assumptions using three different measures of socioeconomic position, allowing for a more comprehensive measure of inequalities in Lithuania and the possible impact of alcohol control policies. Lastly, the data will provide an update to the trajectories of mortality inequalities in Lithuania, which used to be among the largest in Europe.[5]

One of the main limitations of this study is inherent to evaluating effects of public health policies. As these policies are usually not implemented under controlled and randomized conditions, establishing causality for policy effects constitutes a real challenge. Using interrupted time series analyses, an internal control is build from previous observations, which makes this technique one of the most robust methods to evaluate the effects from policy interventions.[30] Acknowledging that the robustness of findings from interrupted time series analyses is heavily depending on controlling for possible confounders, we will attempt to rule out alternative explanations for possible policy effects observed in the time series. Moreover, the results should be interpreted in line of other effective alcohol control policies implemented in Lithuania in close proximity to the 2017 increase of alcohol excise taxation, including sales restrictions in petrol stations, restrictions of opening hours, ban of advertisements, and increasing the minimum legal drinking age.[14]

In conclusion, the results of this study will not only provide an update on the trajectories of mortality inequalities in Lithuania but also offer valuable insights on possible effects of raising alcohol excise taxation on health inequalities. The study findings will be compared against assumptions and estimates from previous modelling studies and are expected to strengthen the evidence base regarding the real-world effectiveness of alcohol control policies.

## Data Availability

Not applicable.

## Authors’ contributions

All authors contributed to conceptualizing the outlined analytical approach. JM wrote the first draft and all co-authors commented on it and provided important intellectual revisions.

## Funding statement

This work was supported by the US-based National Institute of Alcohol Abuse and Alcoholism, grant number 1R01AA028224-01.

## Competing interests statement

None declared.

## Notes

### Competing Interest Statement

The authors have declared no competing interest.

### Author Declarations

The project has been granted research ethics approval 050/2020 by CAMH Research Ethics Board on April 17, 2020, renewed on March 30, 2021.

